# Evaluating Large Reasoning Model Performance on Complex Medical Scenarios In The MMLU-Pro Benchmark

**DOI:** 10.1101/2025.04.07.25325385

**Authors:** R. E. Hoyt, D. Knight, M. Haider, M. Bajwa

## Abstract

Large language models (LLMs) have emerged as a major force in artificial intelligence, demonstrating remarkable capabilities in natural language processing, comprehension, and text and image generation. Recent advancements have led to the development of LLMs specifically designed for medical applications, showcasing their potential to revolutionize healthcare. These models can analyze complex medical scenarios, assist in diagnoses, and provide treatment recommendations. However, evaluating the accuracy and reliability of LLMs in medicine remains a crucial challenge. The output may not be current and could suffer from inaccurate information, known as hallucinations. In early 2025, DeepSeek R1 was released, which is a large reasoning model (LRM) that includes the “chain of thought” reasoning that made it more transparent than any LLM that preceded it. This study utilized the new MMLU-Pro benchmark, which is a more complex Q&A dataset compared to the Massive Multitask Language Understanding (MMLU). DeepSeek R1 was used to analyze the Q&A dataset primarily for accuracy, but medical scenario Q&As are only one facet of a comprehensive assessment. The study found that DeepSeek R1 had an accuracy rate of 96.3% on 162 medical scenarios after reconciliation with subject matter experts on 23 questions. Our findings contribute to the growing body of knowledge on LLM applications in healthcare and provide insights into the strengths and limitations of DeepSeek R1 in this domain. DeepSeek R1 demonstrates excellent accuracy along with unique transparency. Our analysis also highlights the need for multifaceted evaluation methods that go beyond simple accuracy metrics to ensure the safe and effective deployment of LLMs in medical settings.

## Introduction

Large language models (LLMs) are the most common application of generative artificial intelligence (GenAI). LLMs, trained on massive text data, have shown promise in various domains, including medicine, where they can potentially transform healthcare delivery and patient care. Characterized by their immense scale, these models often consist of hundreds of millions to billions of parameters and are trained on vast textual datasets. The advent of multimodal LLMs is bringing about a paradigm shift in the medical field by offering the capability to process and generate diverse data types such as text, images, sounds, and videos. This ability has revolutionized the field of radiology and telemedicine. [1] In the medical field, LLMs are being explored for a wide range of applications, such as diagnosing diseases, providing treatment recommendations, analyzing medical records, acting as ambient scribes, conducting literature reviews, medical education, project management, and answering patient queries. [2] They can also accelerate drug discovery, analyze biomedical literature, and predict the potential function of newly discovered genes. [1] Furthermore, LLMs can streamline clinical workflows, automate documentation, and improve the efficiency of administrative medical processes. [3] The use of GenAI in medicine so far has been mostly related to information retrieval and efficiency and less related to impacting clinical services. [4]

One logical way to evaluate LLM knowledge in the medical domain is to determine how well they answer standard medical questions. The MedQA USMLE is a dataset of multiple-choice medical questions derived from the United States Medical Licensing Examination (USMLE). [5] It consists of 12,723 multiple-choice questions (MCQs) with four possible answers. [5] There is a dynamic leaderboard that keeps scores as newer LLMs are released. As of the time of this submission, OpenAI-o1 scored the highest at 96.5%. [6]

While board review-styled questions are a logical approach to evaluating medical knowledge, some have argued that this approach is inadequate to evaluate medical reasoning. [5]

In addition, the scores for MedQA plateaued in 2024. [7] Therefore, there is a movement to develop different repositories of medical questions and answers to improve on MedQA questions. [7] For example, MetaMedQA is a modified question test bank that tests metacognition by removing some answers and adding misleading and fictitious answers to make testing more difficult. [7] Results indicated that twelve LLMs did well when the correct answer was removed but did extremely poorly with misleading or fictitious scenarios. [8]

Massive Multitask Language Understanding (MMLU) is another large question-and-answer dataset developed by a team of researchers in 2020. It consists of approximately 16,000 multiple-choice questions spanning 57 academic subjects. These subjects cover a wide range of knowledge domains, including humanities, social sciences, STEM, and professional fields such as law and health. [9] LLMs have been tested on this question bank, with a leaderboard keeping a score of their performance. [10] Recently, work at the Tiger Lab, which is part of the University of Waterloo, developed the MMLU-Pro modification. [11-12] MLLU-Pro was distilled down to 14 categories covering the major domains and includes approximately 12,000 questions. The questions are more complex and more reasoning-focused. It expands the MCQ answer options from four to up to ten, reducing the likelihood of random guessing and increasing the evaluation’s complexity. MMLU-Pro also eliminates trivial and noisy questions found in the original MMLU, making it a more discriminative benchmark. There is also an MMLU-Pro leaderboard to track accuracy by newly released LLMs. [13]

LLMs have steadily improved since 2022, as evidenced by the improving scores on multiple leaderboards. In early 2025, several “reasoning” LLMs were released that use “chain of thought” (CoT) reasoning, which means they output an answer using logical steps, like how a human thinks. Some refer to these new LLMs as large reasoning models or LRMs. Through multiple training steps, the model is able to self-correct, reevaluate, and refine its opinion through CoT. DeepSeek R1, developed in China, is a 671-billion-parameter state-of-the-art open-source LLM released in late January 2025. [15-16] DeepSeek R1 is unique as it utilizes supervised fine-tuning, reinforcement learning, and a Mixture of Experts (MoE) methodology. An MoE is a machine learning approach that divides an AI model into sub-networks, or “experts,” each specializing in a subset of the input data to perform a task. The tasks are activated sequentially and iteratively with collaboration among the experts. [15-16] Unlike non-reasoning models, zero-shot prompting with clear but brief instructions is best. [17] DeepSeek R1 was less expensive to develop, and the input and output inference costs are low. [18] There are concerns about privacy and security with any platform developed outside of the US, but because it is open-source, it can be hosted on a local computer in addition to access via an API. The latency, or time to generate an answer, is slower due to the time it takes to complete the CoT steps. [19] DeepSeek R1 scores on common benchmarks are as follows: MedQA (94.3), MMLU (90.8), and MMLU-Pro (84). There is no score for the sub-category “health,” however, so the score of 84 on MMLU-Pro is an overall score and self-reported. [17] [20]

To date, there are no studies reporting the performance of DeepSeek on medical scenarios that require reasoning in addition to medical knowledge. The goal of this paper is to study the performance of DeepSeek R1 on complex medical scenarios found within the MMLU-Pro benchmark.

## Methods

The MMLU-Pro databank, consisting of about 12000 questions and answers in 14 categories, was downloaded from the hosting website using Python programming. [10] The health category was selected by filtering, and this category consisted of the following subcategories: virology, clinical knowledge, anatomy, medical genetics, nutrition, college medicine, aging, and professional medicine. We further filtered this category to only include the professional medicine subcategory, which included 166 complex medical scenarios. Questions were further categorized into twenty-one medical specialties. (See Table 1). Approximately a third of the questions were related to adult or pediatric primary care. About 15% were emergency room visits, and the remainder were divided among multiple subspecialties.

**Table 1.** Specialty and number of medical scenario questions.

| Specialty | Questions |
| --- | --- |
| Primary care | 37 |
| ER medicine | 24 |
| Pediatrics | 16 |
| OB-Gyn | 11 |
| Neurology | 10 |
| Statistics | 9 |
| Infectious disease | 7 |
| Endocrinology | 6 |
| General surgery,<br>Gastroenterology,<br>Cardiology | 5 |
| Oncology | 4 |
| Nephrology, Genetics,<br>Psychiatry | 3 |
| Immunology/Allergy,<br>Trauma surgery,<br>Rheumatology,<br>Vascular surgery,<br>Orthopedics,<br>Pulmonology | 2 |

We deleted duplicate questions and four questions that had formatting errors. A prompt was created: “As *a medical consultant, you will respond to questions about various medical scenarios. This is a multiple-choice question with up to 10 possibilities. Select the answer that is most likely and estimate your degree of confidence from 1-5, where 5 is very confident. Report the correct answer A-J*.”

Following Griot et al. [8], a confidence level was added as: High confidence (4-5): >90% alignment with diagnostic criteria, medium confidence (2-3): 70-90% alignment and low confidence (1): <70% certainty or conflicting indicators.

Despite the statement that MMLU-Pro frequently has a maximum of 10 possible answers, it was not uncommon to have as few as four answers.

A well-known AI search engine platform hosted an uncensored version of DeepSeek. [21-22] We inputted the prompt, then the question, and finally the answer in a single paragraph. DeepSeek generated the following steps: 1. It first performed a literature review for most questions. 2. It began a very iterative process that consisted of multiple steps as part of chain-of-thought reasoning. 3. It then arrived with a letter answer and a confidence level. Lastly, the answer was followed by an explanation of why it was chosen and why it excluded the other potential answers, supported by literature citations. (see example of DeepSeek R1 workflow in Appendix A.) In those cases of disagreement with the stated answers, the question/answer pairs were uploaded to Claude 3.7 Sonnet Reasoning as a second opinion. [26] When DeepSeek produced a different answer than the answer provided by MMLU-Pro, the scenario was evaluated by subject matter experts (SMEs) from the Mayo Clinic and two women’s clinics in a blinded fashion where they were given the scenario question but not the answer.

## Results

One hundred and sixty-two question/answer pairs were analyzed with DeepSeek R1. There was an initial agreement in 85.8% of the cases between the stated MMLU-Pro answers and DeepSeek. There were 23 instances of disagreement, but in 19 instances, Claude 3.7 Sonnet Reasoning agreed with DeepSeek’s opinion. In three instances, Claude 3.7 Sonnet Reasoning agreed with the stated answer; in one instance, it disagreed with the stated answer and the one given by DeepSeek. In the 23 instances of disagreement, the results were reconciled by an SME. The results are displayed in Table 2.

**Table 2.** DeepSeek Incorrect Answers and Resolution.

| DeepSeek Incorrect Answers | LRM Second Opinion | SME Opinion | Final Resolution |
| --- | --- | --- | --- |
| 23 scenarios | 19 Concurrence | 15 times opinion agreed with DeepSeek<br>8 times opinion disagreed with DeepSeek | 6 incorrect answers, confirmed by SMEs.<br>Accuracy = 96.3% |

After reconciliation by the SMEs, the accuracy rate increased to 96.3% or 6 incorrect answers out of a total of 162 questions. The confidence rating was almost exclusively at the 5 level, indicating complete confidence in the answers. In twelve instances, there was a rating of 4, and there was no correlation with those answers that disagreed with the MMLU-Pro answers. A confidence level of 4 was the lowest confidence score recorded in this evaluation.

On a random sample of 50 medical scenarios, the minimum, maximum, average, and range were calculated for references, unrelated references, and reasoning steps. Table 3 presents the results of these calculations. Each reasoning step was expandable and could vary from a few short sentences to a very lengthy paragraph.

**Table 3.** References, unrelated references, and reasoning step calculations.

|  | Minimum | Maximum | Average | Range |
| --- | --- | --- | --- | --- |
| References | 0 | 46 | 16.04 | 46 |
| Unrelated references | 0 | 11 | 4.82 | 11 |
| Reasoning steps | 3 | 56 | 12.16 | 53 |

It is not known why DeepSeek chose not to cite any references on 11 of the 50 medical scenarios. The lack of references did not correlate with incorrect answers or reasoning steps. The majority of references were from well-known journals, healthcare organizations such as the Mayo Clinic, and clinical practice guidelines. We did not find any “hallucinated references,” meaning that they were made up and did not exist. The references were hyperlinked and not static, making it easy to review and confirm. The unrelated references were not germane to the medical scenario and occasionally consisted of non-medical references. The average number of “unrelated references” was 4.82 with a range of 0-11. Fourteen questions were associated with no unrelated references. It was not obvious why the reasoning steps varied widely, and the steps taken did not correlate with the confidence level or the degree of correctness.

## Discussion

DeepSeek R1 is a unique LRM that functions well and answers complex medical scenarios on MMLU-Pro. While we have the overall scores for other LLMs in the MMLU-Pro “health” category, we don’t have scores for DeepSeek R1. It is, therefore, not known if DeepSeek R1 would have scored as well for all subcategories under the health category.

DeepSeek is one of five popular LRMs: DeepSeek R1, OpenAI o3 mini, Claude 3.7 Sonnet Reasoning, Gemini 2.5 Pro, and QwQ 32B. [16, 27-30] All display reasoning steps, but QwQ 32B and Gemini 2.5 Pro do not display a literature search, so they are less transparent. Despite the differences in transparency and resources, the non-DeepSeek LRMs informally tested came to the same conclusions when queried with a medical scenario. Therefore, at this point, we don’t know how the other LRMs compare with DeepSeek R1 without formal testing. DeepSeek R1 has the longest latency period because it spends more time gathering resources and outlining reasoning steps. This assumption is further supported by a formal analysis that revealed that the outputted tokens per second are only 23 for DeepSeek and 215 for Gemini 2.0 Flash. [18] This output delay does not appear to have clinical implications for the average user.

There is concern about DeepSeek’s security. DeepSeek failed security checks and has not reported any safety evaluations, raising uncertainties about the trustworthiness of the LRM. [31]

Importantly, implementing any AI in a hospital or healthcare educational setting requires a robust infrastructure, staff training, and maintenance. There needs to be regular model updates to stay current, which is an additional expense. Furthermore, hallucinations and bias need to be addressed effectively. [1]

Our analysis of MMLU-Pro as a question and answer bank found that it contained multiple duplicates just in the professional medicine subcategory. In addition, there were questions where the stated answer was incorrect. It is likely that other medical MCQ banks contain similar errors.

There are issues that can make MedQA-style MCQs less useful, like the “cueing effect” or answer choices that give test-takers subtle hints that help them get the right answer without fully understanding the concepts behind the questions. [32] Another limitation of MCQs is the phenomenon of “testwiseness.” Testwiseness refers to the ability of test-takers to use strategies such as eliminating implausible options or identifying clues in the question to determine the correct answer. [33] MCQs may also fail to assess a test taker’s ability to understand and apply knowledge in complex, real-world situations. Clinicians not only have to have the correct diagnosis, but they also need to appreciate the timing of when to treat or not to treat. Many MCQs are designed to test factual knowledge rather than the ability to reason through complex clinical scenarios. [34] Lastly, only 5% of studies evaluating LLMs used real patient data. [35]

The questions in the professional medicine subcategory covered twenty-one specialties, so it is likely that DeepSeek R1 would outscore most physicians. Even when tested in their specialty, physicians may not outperform frontier LLMs. In a study published in 2025, internal medicine residents were quizzed on questions from a commercial internal medicine board review test bank, and the average score was lower than any LLM tested. [36]

### Limitations

Importantly, we only evaluated one LRM and one medical question-and-answer set. Most specialties had fewer than 15 specialty-related questions, so this set was an inadequate test for many specialties. The results would likely be different for other LLMs or LRMs and for other question-and-answer datasets. We also did not compare a reasoning model with a biomedical LLM, like MedPaLM-2. [37] We wrote our prompt concisely so that DeepSeek could only respond to the listed answers. This method may not be the best approach, as different prompting strategies might produce different results.

### Future Recommendations

It is unclear how LRMs, like DeepSeek R1, should be used or implemented in clinical medicine. Medical decision-making requires a high level of real-time expertise and experience that may not be achievable by LLMs or LRMs. It is critical to research LLMs using robust research methods before implementation in medical practice. [2] However, despite recognized LLM shortcomings, there is evidence that the average clinician is already using AI, particularly LLMs. A 2024 American Medical Association (AMA) survey found that about three in five (66%) physicians currently use AI in their practice, a significant increase from 38% in 2023. [38]

The AMA’s recent principles for AI in medicine highlight the need for a comprehensive evaluation system for LLMs in medical scenarios used for education and training. [39] This evaluation system should assess not only accuracy and efficiency but also safety, applicability, and impact on patients. It requires collaboration among medical experts, computer scientists, regulatory bodies, and the public to ensure LLMs are used responsibly and effectively. The research should include:

- In-depth analyses [40]
- Faithfulness or how LLM outputs align with confirmed medical knowledge [41]
- Comprehensiveness or the ability of LLMs to capture all aspects of complex medical scenarios [41
- Generalizability or the ability to perform consistently across a variety of scenarios [42]
- Robustness or the LLM’s resilience to perform well regardless of the input data, prompt used, or other factors. [42]

We need better medical benchmarks that utilize real patient data and mirror the thought processes of clinicians. For example, MedHELM is a Stanford University initiative that evaluates LLMs on real electronic health record data. Evaluation categories include clinical decision support, clinical note generation, patient communication and education, medical research assistance, and administration and workflow. [43] Another recent guideline is the human-oriented QUEST (Quality of Information, Understanding and Reasoning, Expression Style and Persona, Safety and Harm, and Trust and Confidence), intended to evaluate LLMs in healthcare. [44]

In the future, we plan to evaluate the same 162 medical scenarios with DeepSeek R1 but not provide the answer to see how it performs in the absence of the cueing effect. This approach should be a more meaningful evaluation of how LRMs analyze medical scenarios without answer choices.

## Conclusions

LRMs, like DeepSeek, are able to effectively answer questions related to complex medical scenarios in multiple subspecialties. Further research is needed to determine how they should be leveraged in clinical medicine. Although further research is needed and warranted, these newest models are so impressive that it is likely that they will be used despite warnings to limit use until further research is completed.

## Data Availability

All data produced in the present study are available upon reasonable request to the authors

## Acknowledgements

**Subject Matter Experts**

Thanks to the following individuals for their expert opinions

Mayo Clinic, Jacksonville, FL

1. Hasadsri, Linda, MD, PhD
2. Sledge, Hanna J
3. Alvarez, Salvador, MD
4. Olomu, Osarenoma U, MD
5. Hodge, David O
6. Shair, Kamal A, MD
7. Muthusamy, Karthik, M.B.B.S.
8. Yataco, Maria L, MD
9. Wysokinska, Ewa M, MD

Capital’s Women Care, Silver Spring, MD

1. Lizardo, Randolph D, MD

Nova Group For Women, Fairfax, VA

1. Andersen, Glenna, MD

**Note:** Quillbot was used to detect grammatical and syntax errors

## Appendix Supplement Material

### Prompt, Question, and Answer

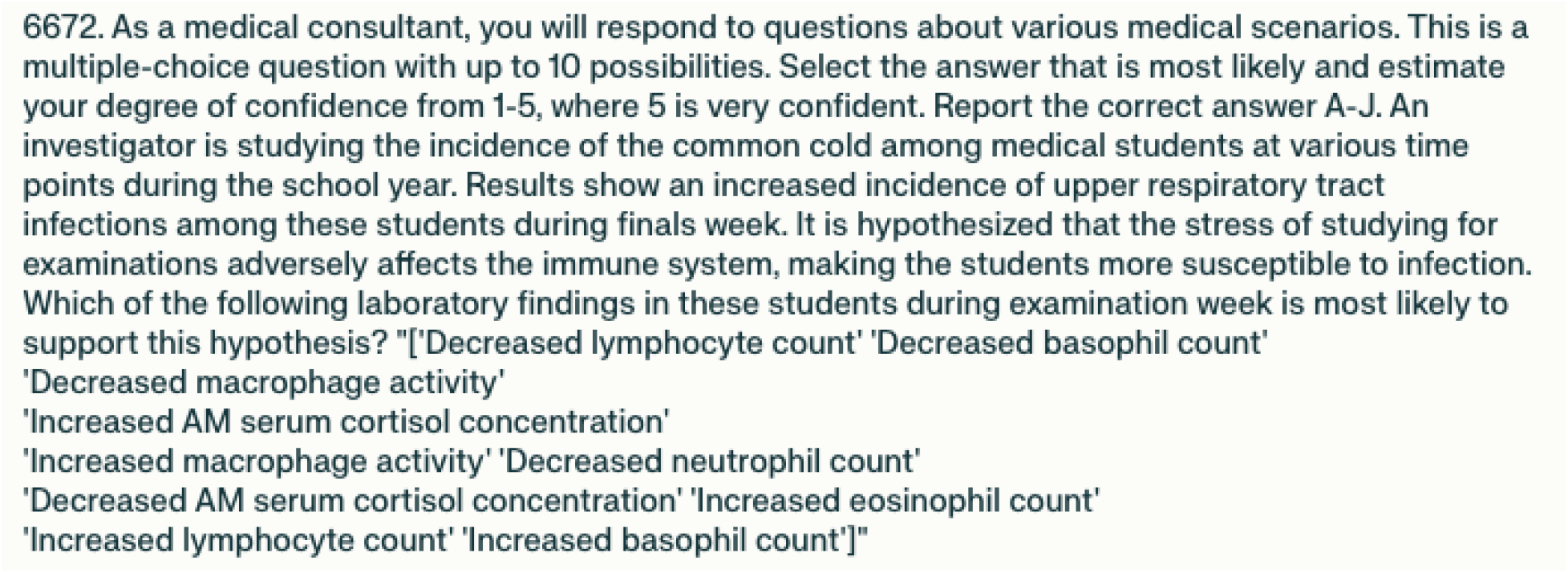

### Literature Search

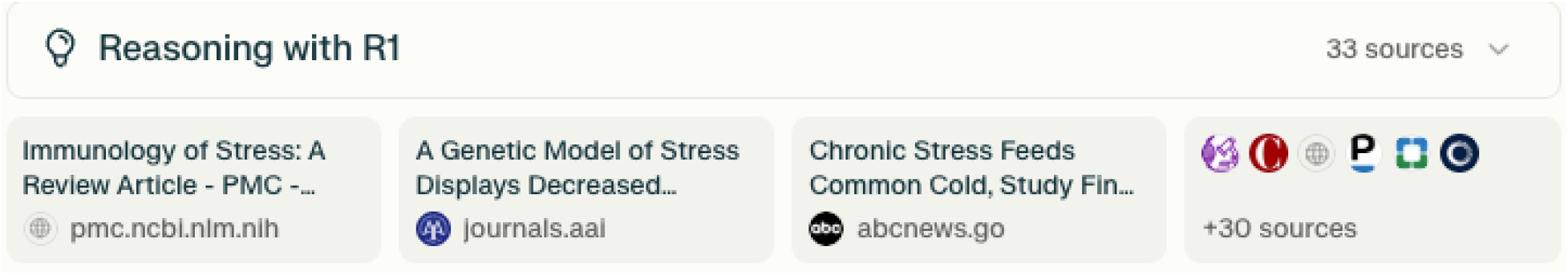

### Reasoning Steps

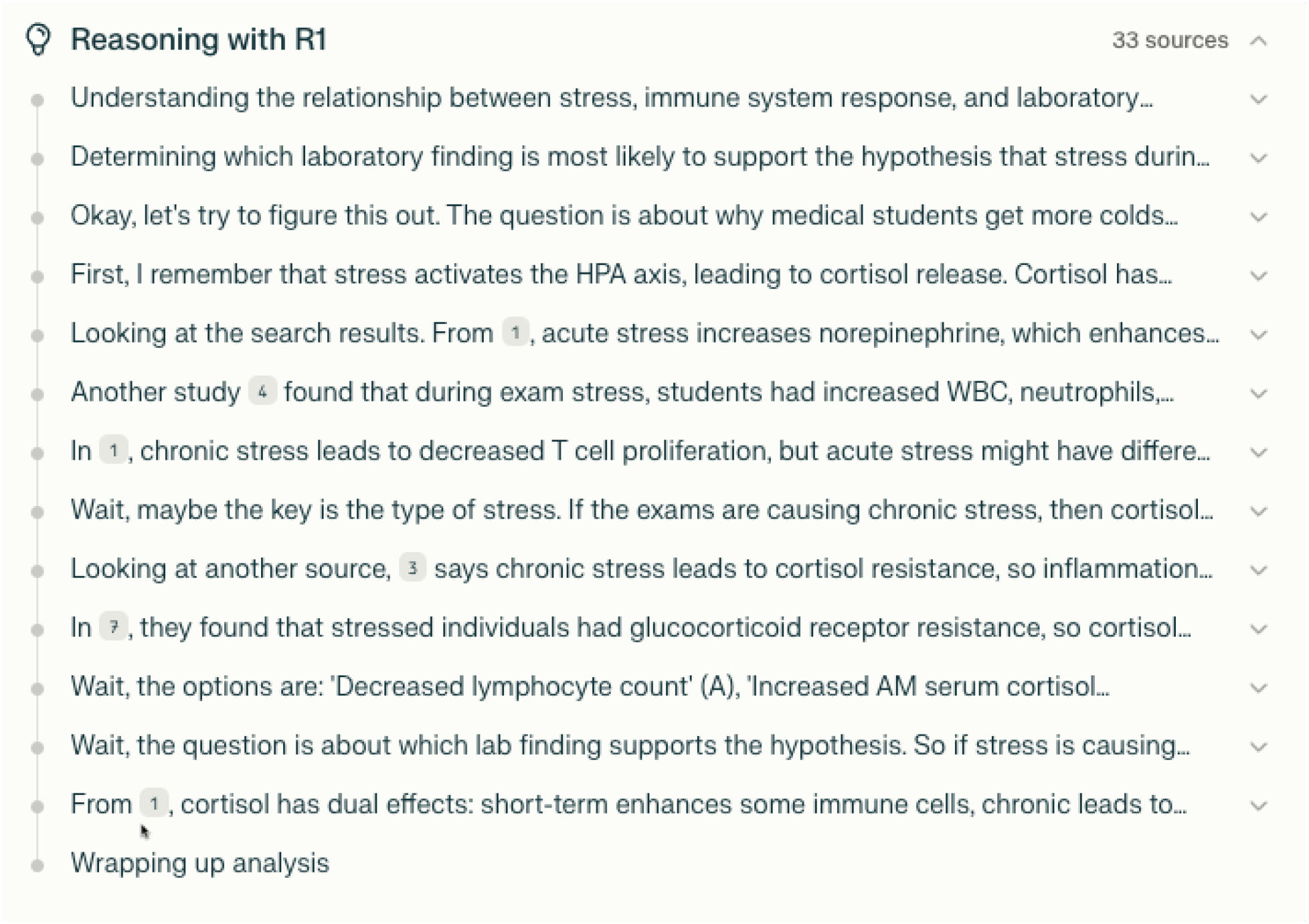

### Answer, Logic, and Conclusion

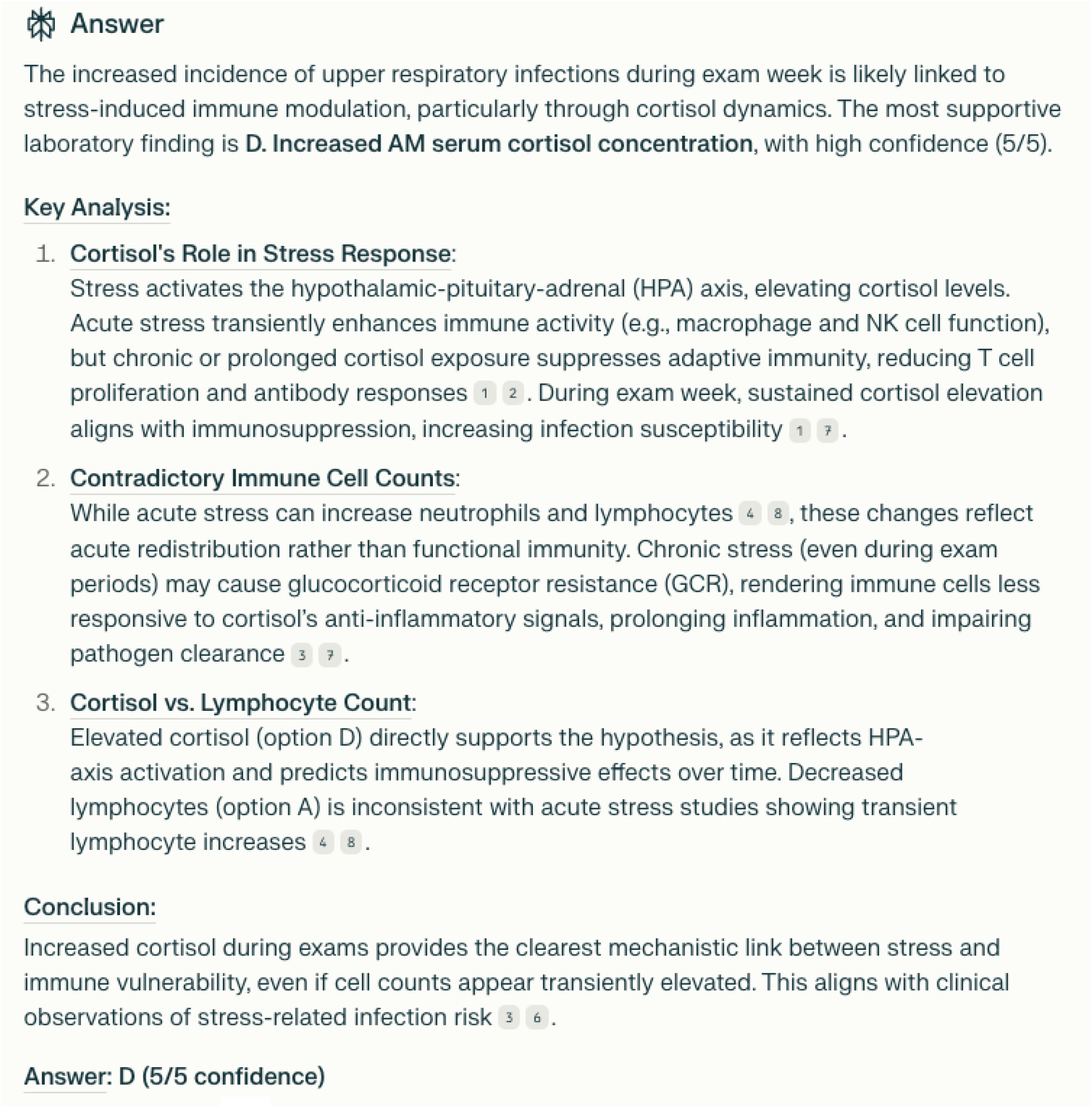

